# The Link Between GABA Levels and P300 Abnormalities in Schizophrenia Spectrum Disorders: Regional and Symptom-Based Insights

**DOI:** 10.1101/2025.03.19.25324186

**Authors:** Berkhan Karslı, Verena Meisinger, Genc Hasanaj, Marcel S. Kallweit, Fanny Dengl, Gizem Vural, Julian Melcher, Maxim Korman, Nicole Klimas, Susanne Schmölz, Antonia Šušnjar, Alexandra Hisch, Lenka Krčmář, CDP Working Group, Emanuel Boudriot, Joanna Moussiopoulou, Vladislav Yakimov, Oliver Pogarell, Andrea Schmitt, Peter Falkai, Thomas Geyer, Lukas Roell, Elias Wagner, Florian J. Raabe, Daniel Keeser

## Abstract

According to the excitation-inhibition imbalance theory, GABAergic and glutamatergic systems influence the clinical symptoms and particularly cognitive deficits in schizophrenia spectrum disorders (SSD). These systems have been found disrupted in the anterior cingulate cortex (ACC) and dorsolateral prefrontal cortex (DLPFC) in SSD, and are associated with P300 abnormalities in electroencephalography recordings. Therefore, we explored the relationships among MRS-derived GABA and Glx levels in the ACC and left DLPFC (lDLPFC), auditory P300 amplitudes and latencies, cognition, and symptom severity in SSD. We grouped patients into higher (SSD+) and lower (SSD−) symptom severity clusters based on PANSS total scores. P300 amplitudes were lower in SSD patients than healthy controls at central and parietal sites. SSD+ exhibited widespread reductions at frontal, central, and parietal areas, while SSD− showed reductions limited to the parietal region. GABA levels in the lDLPFC were higher in SSD− compared to controls and were positively associated with P300 amplitudes at central and parietal sites within SSD- and overall SSD group. Although P300 amplitudes positively correlated with the BACS composite scores and behavioral performance, lDLPFC GABA levels showed no direct association with cognitive or behavioral performance. ACC GABA, ACC Glx, and lDLPFC Glx levels showed no group differences or P300 associations. Our findings suggest P300 amplitude reductions as a marker of cognitive dysfunction in SSD, more pronounced in patients with higher disease severity, and that enhanced lDLPFC GABA may help offset these reductions. Our work provides the first empirical evidence of the interplay between the GABAergic system and cortical electrophysiological signal patterns mirroring cognitive dysfunction in SSD.

## Introduction

Schizophrenia spectrum disorders (SSD) are complex neuropsychiatric disorders characterized by positive symptoms such as hallucinations and delusions, negative symptoms such as anhedonia and avolition, and cognitive impairment^1^. Cognitive deficits, which are directly correlated with patients’ daily functioning, remain largely unresponsive to current treatments that primarily target dopamine D2 receptors^2^, perhaps due to a limited understanding of the underlying mechanisms. One of the most consistently replicated neural correlates of SSD-related cognitive impairments is P300 amplitude reduction^3,4^. P300 is an event-related potential (ERP) component, typically elicited as a positive peak around 300 ms after presenting an infrequent target stimulus within a series of standard stimuli in an oddball task, was shown to reflect a cognitive process^5^. The amplitude of the P300 component of the event-related potential (ERP) is well-established for its sensitivity for target probability^6^. This sensitivity may indicate the ability to keep track of event probabilities, or statistical environmental structure, to facilitate perceptual decision-making in the future^7^. Moreover, P300 amplitude reductions in SSD are more pronounced with higher clinical symptom severity and fluctuate with longitudinal changes in symptom levels, becoming greater during symptom deterioration and milder when symptoms improve^8,9^.

Neurochemical imbalances, especially disruptions in the excitation-inhibition (E/I) balance mediated by glutamate and gamma-aminobutyric acid (GABA), have been suggested to play a crucial role in the pathophysiology of SSD^10^. Such disruptions can lead to neural circuit dysfunctions contributing to cognitive deficits^5^. Magnetic resonance spectroscopy (MRS) studies comparing GABA and glutamate levels in individuals with SSD and healthy controls have yielded inconclusive findings across brain regions and patient characteristics^11,12^. Symptom heterogeneity across patient groups or region-specific E/I balance differences may be an explanation for these inconsistencies, which we have attempted to address in our study. Another factor that may contribute to these inconsistencies is the relatively small sample sizes in many MRS studies; our study aims to address this issue by including a comparatively larger sample. Further supporting the role of neurotransmitter imbalances in SSD, pharmacological studies administering N-methyl-D-aspartate receptor (NMDAR) antagonists and GABA agonists to healthy individuals show that disruptions in glutamatergic and GABAergic neurotransmission reduce P300 amplitude, mirroring changes seen in SSD, and suggesting a role for these systems in P300 abnormalities^5^.

The dorsolateral prefrontal cortex (DLPFC) and anterior cingulate cortex (ACC) are critical regions implicated in cognitive dysfunctions in SSD. Structural changes, including gray and white matter reductions, and functional changes are reported in both regions^13,14^. The ACC and DLPFC are crucial for cognitive control and higher-order functions^15–17^. In SSD, reduced activation in the ACC and DLPFC disrupts cognitive functions like monitoring, decision-making, and working memory^18^, and is involved in P300 generation via their connection to the intraparietal sulcus^19^. In patients with SSD, ACC activation during the P300 component is reduced, indicating deficits in top-down regulation of stimulus relevance^20^. These frontal P300 reductions are hypothesized to impair response inhibition or initiation^21^. These findings highlight the DLPFC and ACC as key regions implicated in P300 abnormalities in SSD.

The present study aims to investigate the role of the GABAergic and glutamatergic systems in P300 abnormalities and related cognitive deficits in SSD, while accounting for symptom heterogeneity. Although several neurotransmitter systems have been implicated in P300 modulation in SSD^5^, to our knowledge, no study has directly linked brain metabolite levels with this ERP component. First, we sought to replicate the well-established P300 amplitude reductions in patients. Second, to account for symptom heterogeneity and its previously reported association with P300 amplitude reductions^8,9^, we clustered patients based on symptom severity, hypothesizing that more symptomatic patients would show greater reductions. Third, we examined whether GABA and Glx (a composite of glutamate and glutamine) levels in the ACC and DLPFC differ between healthy controls, the overall patient group, and patient clusters. Fourth, we examined whether these metabolite levels are associated with P300 amplitude reductions in the overall patient group and in patient clusters. Finally, we examined how metabolite levels and disrupted P300 indices are each related to cognition and behavioral task performance in the patient group and clusters. These findings may help clarify the neurochemical contributions to P300 abnormalities and their impact on cognition in SSD.

## Methods

### Study Sample and Design

All 214 participants (107 patients with Schizophrenia Spectrum Disorder [SSD] and 107 healthy controls [HC]) included in the study were recruited as part of the Clinical Deep Phenotyping (CDP) study^22^, a project within the Munich Mental Health Biobank^23^ (see Table 1). The study, approved by the Ethics Committee of the Faculty of Medicine, LMU Munich (project numbers: 20-0528 and 22-0035) and registered in the German Clinical Trials Register (DRKS; registration ID: DRKS00024177), included individuals aged 18–65 years recruited from the in- and outpatient population of the Department of Psychiatry and Psychotherapy at the LMU University Hospital, along with healthy controls. The majority of patients were treated with second generation antipsychotic medications. Inclusion criteria for patients were a diagnosis of schizophrenia (SZ), schizoaffective disorder (SZA), brief psychotic disorder (BrPsyD), or delusional disorder (DD) according to the MINI International Neuropsychiatric Interview^24^. General exclusion criteria included any primary psychiatric disorder other than those listed, clinically relevant central nervous system (CNS) disorders (past or present), pregnancy, and major language barriers. Symptom severity was assessed by trained staff using the Positive and Negative Syndrome Scale (PANSS)^25^. Cognitive function was assessed using the Brief Assessment of Cognition in Schizophrenia (BACS)^26^, and z-standardized BACS composite scores were calculated. All participants completed P300 and MRS assessments in separate sessions. Detailed information about the assessments and score calculations is included in the Supplemental Methods.

**Table 1.** Sample characteristics table. HC = healthy control participant, SSD = schizophrenia spectrum disorder, BACS = Brief Assessment of Cognition in Schizophrenia, BMI = body mass index, CPZeq = chlorpromazine equivalent dose, GAF = Global Assessment of Functioning, PANSS = Positive and Negative Syndrome Scale. *^a^* Fisher’s exact test, *^b^* Welch’s t test.

|  | SSD |  | HC |  | p |
| --- | --- | --- | --- | --- | --- |
| | Mean ± SD or n (%) | n | Mean ± SD or n (%) | n | |
| **Demographic Characteristics** |  |  |  |  |  |
| Age, Years | 38.66 ± 12.25 | 107 | 33.93 ± 12.22 | 107 | 0.005<sup>b</sup> |
| Sex, Female:Male (% Female) | 32:75 (29.9%) |  | 55:52 (51.4%) |  | 0.002<sup>a</sup> |
| BMI | 28.44 ± 5.66 | 105 | 23.42 ± 3.35 | 105 | < 0.001<sup>b</sup> |
| Smoking, Yes:No (% Yes) | 53:52 (50.5%) |  | 8:95 (7.8%) |  | < 0.001<sup>a</sup> |
| **Comorbidities** |  |  |  |  |  |
| Diabetes, Yes:No (% Yes) | 4:103 (3.7%) |  | 1:106 (0.9%) |  | 0.369<sup>a</sup> |
| Hypertension, Yes:No (% Yes) | 17:90 (15.9%) |  | 4:103 (3.7%) |  | 0.005<sup>a</sup> |
| **Disease Characteristics** |  |  |  |  |  |
| Disease Duration, Months | 142.63 ± 121.92 | 105 | | | |
| Antipsychotic Treatment Duration, Months | 123.62 ± 112.76 | 95 | | | |
| Benzodiazepine Use, Yes:No (% Yes) | 13:94 (12.1%) |  |  |  |  |
| CPZeq, mg | 343.22 ± 249.21 | 104 | | | |
| Remission Andreasen, Yes:No (% Yes) | 51:55 (48.1%) |  |  |  |  |
| PANSS Positive Symptoms | 12.64 ± 5.28 | 106 | 7.26 ± 0.78 | 107 | < 0.001<sup>b</sup> |
| PANSS Negative Symptoms | 13.42 ± 5.71 | 106 | 7.42 ± 0.97 | 107 | < 0.001<sup>b</sup> |
| PANSS General Symptoms | 28.97 ± 8.9 | 106 | 17.06 ± 1.55 | 107 | < 0.001<sup>b</sup> |
| PANSS Total Score | 55.03 ± 17.3 | 106 | 31.74 ± 2.64 | 107 | < 0.001<sup>b</sup> |
| GAF | 53.42 ± 11.37 | 104 | 89.9 ± 6.49 | 107 | < 0.001<sup>b</sup> |
| BACS Composite z Score | -1.84 ± 1.63 | 99 | 0 ± 1 | 104 | < 0.001<sup>b</sup> |
| **Diagnosis** |  |  |  |  |  |
| Schizophrenia | 67 (62.6%) |  |  |  |  |
| Schizoaffective Disorder | 30 (28.0%) |  |  |  |  |
| Brief Psychotic Disorder | 5 (4.7%) |  |  |  |  |
| Unspecified SSD | 4 (3.7%) |  |  |  |  |
| Delusional Disorder | 1 (0.9%) |  |  |  |  |

### P300 Recording and Preprocessing

The P300 was recorded using an Electro-Cap (Electro-Cap International, Inc, Eaton, OH, USA) following the International 10-20 system, connected to a 32-channel BrainAmp amplifier (Brain Products, Martinsried, Germany) in a quiet, dimly lit cabin at the EEG department at the Department of Psychiatry and Psychotherapy, University Hospital LMU Munich. Participants performed an auditory oddball task wearing Phillips headphones, pressing a button in response to infrequent target tones (20% of 700 trials) randomly placed among standard tones (80% of trials) while keeping their eyes closed for 18 minutes.

EEG data were processed following a pipeline adapted from Adams et al.^27^ using MATLAB (The Mathworks Inc.) with EEGLAB (v2022.0)^28^. Signals were re-referenced, filtered, segmented, and subjected to multi-step artifact rejection (including ICA-based removal) before channel interpolation and baseline correction. Detailed thresholds for epoch/channel exclusion and other preprocessing steps are described in the Supplemental Methods.

For P300 analysis, ERPs were averaged by tone condition at each electrode. Region averages were computed for three regions: Frontal (“FP1”, “FP2”, “F3”, “F4”, “Fz”, “F7”, “F8”, “FC1”, “FC2”, “FC5”, “FC6”), Central (“C3”, “C4”, “Cz”), and Parietal (“P3”, “P4”, “Pz”). Regions were defined to enhance interpretability, while results for midline electrodes (“Fz”, “Cz”, “Pz”) were separately reported in the Supplemental Results. The averaged ERPs were subsequently bandpass filtered (0.05–30 Hz, IIR filter). For P300 component analysis, latency and amplitude were extracted from the filtered averaged ERP waveforms at each cluster. Peaks were identified within the 250–400 ms window using a positive peak detection algorithm (get_peak) from MNE-Python (v1.8.0)^29^, capturing the latency and amplitude of the most positive value within the specified time window. Amplitude and latencies for the target tone were used for further analysis.

### MRI Acquisition and Preprocessing

All participants were scanned on a Siemens 3T Magnetom Prisma scanner at the Neuroimaging Core Unit Munich (NICUM) using a 32-channel head coil. T1-weighted images were acquired with a magnetization-prepared rapid acquisition gradient echo sequence (slice thickness: 0.8 mm, repetition time: 2500 ms, echo time: 2.22 ms, flip angle: 8°, FoV: 256 mm^2^).

For single-voxel proton magnetic resonance spectroscopy (¹H-MRS), trained MRI operators manually placed a 20 × 20 × 20 mm^3^ volume of interest (VOI) in the anterior cingulate cortex (ACC; for 58 HC and 63 SSD participants) or a 30 × 30 × 15 mm^3^ VOI in the left dorsolateral prefrontal cortex (lDLPFC; for 53 HC and 50 SSD participants), using the T1-w images for anatomical guidance. For both regions, VOI placement used a template reference image and defined anatomical landmarks. A fastestmap sequence was used for B0 shimming, followed by a MEGA-semi-localized by adiabatic selective refocusing (MEGA-sLASER) sequence (repetition time: 3000 ms, echo time: 68 ms)^30^ for MRS acquisition. Each scan included a water-suppressed acquisition (128 averages) and a non-water-suppressed acquisition (8 averages), used as a reference to correct the baseline and improve background noise estimation.

MRS data were processed by using Osprey (v2.5.0)^31^ and LCModel (Linear Combination Model, v6.3-1R)^32^. The Osprey Processing Module was used to optimize raw MRS data for analysis, voxel registration, and tissue segmentation. Voxel placement consistency was checked visually using the generated coregistration images. All processed spectra were subsequently analyzed in LCModel, using basis sets specifically tailored for this sequence at the NICUM Siemens 3T Magnetom Prisma scanner. Glx was derived from the OFF spectrum, and GABA from the DIFF spectrum. Finally, metabolite concentrations (millimolar, mM) were corrected for cerebrospinal fluid (CSF) contribution within the volume of interest (VOI) using individual T1w MRI segmentation values derived from the Osprey analysis.

### Statistical Analysis

All statistical analyses were conducted using R (v4.4.3), with significance set at uncorrected p < .05 due to the exploratory nature of this study. Age and sex were included as covariates in all analyses. 95% confidence intervals (CI) were reported for all estimates.

Metabolite concentration values were subjected to quality control measures. Values with Cramer-Rao lower bounds (CRLB) exceeding 30% or signal-to-noise ratios (SNR) less than 5 were replaced with missing values (NA). Subsequently, major outliers, values falling more than three times the interquartile range below the first quartile or above the third quartile, were identified^33^ and also replaced with NA.

All group differences were assessed using analysis of covariance (ANCOVA). For comparisons between two groups (e.g., SSD vs. HC), ANCOVA models included group as the between-subjects factor and age and sex as covariates. For comparisons involving three groups (HC, SSD−, SSD+), multilevel group ANCOVAs were performed. Post hoc pairwise comparisons were conducted using estimated marginal means (EMMs) contrasts. These contrasts allowed for adjusted comparisons between specific groups while controlling for the covariates. Cohen’s d was used as an estimate of the effect size for these contrasts.

Linear regression analyses were used to explore the relationships between metabolite levels and P300 measures. Models included P300 measures as the dependent variable and metabolite levels as predictors, along with the interaction between metabolite levels and group. Age and sex were included as covariates. To obtain robust estimates of the regression coefficients and associated p-values, bootstrapping methods were employed. Specifically, 10000 bootstrap samples were generated for each model using residual resampling. 95% confidence intervals were computed from the bootstrap distributions, and p-values were calculated by inverting the confidence intervals using boot.pval library (v0.7.0) in R^34,35^.

Partial correlation analyses were conducted to examine the relationships of metabolite levels and P300 measures with behavioral and cognitive measures, while controlling for age and sex. This method allowed for the assessment of the strength of associations between variables independent of the covariates.

To capture the symptom heterogeneity underlying P300 amplitude fluctuations within the patient group, k-means clustering (100 runs with random starting points, selecting the solution with minimal within-cluster variances) was performed based on PANSS total scores. Based on multiple clustering solutions evaluated with silhouette scores, a two-cluster solution was selected as the optimal solution (silhouette score = 0.62). This choice also reflects our aim to form the most clinically distinct groups characterized by symptom severity, while maintaining reasonable group sizes. Participants were thus assigned to one of two clusters: SSD− (lower-symptom) or SSD+ (higher-symptom), which were used in subsequent analyses.

For a summary of acquisition and analysis steps, please refer to Figure 1.

**Figure 1.**
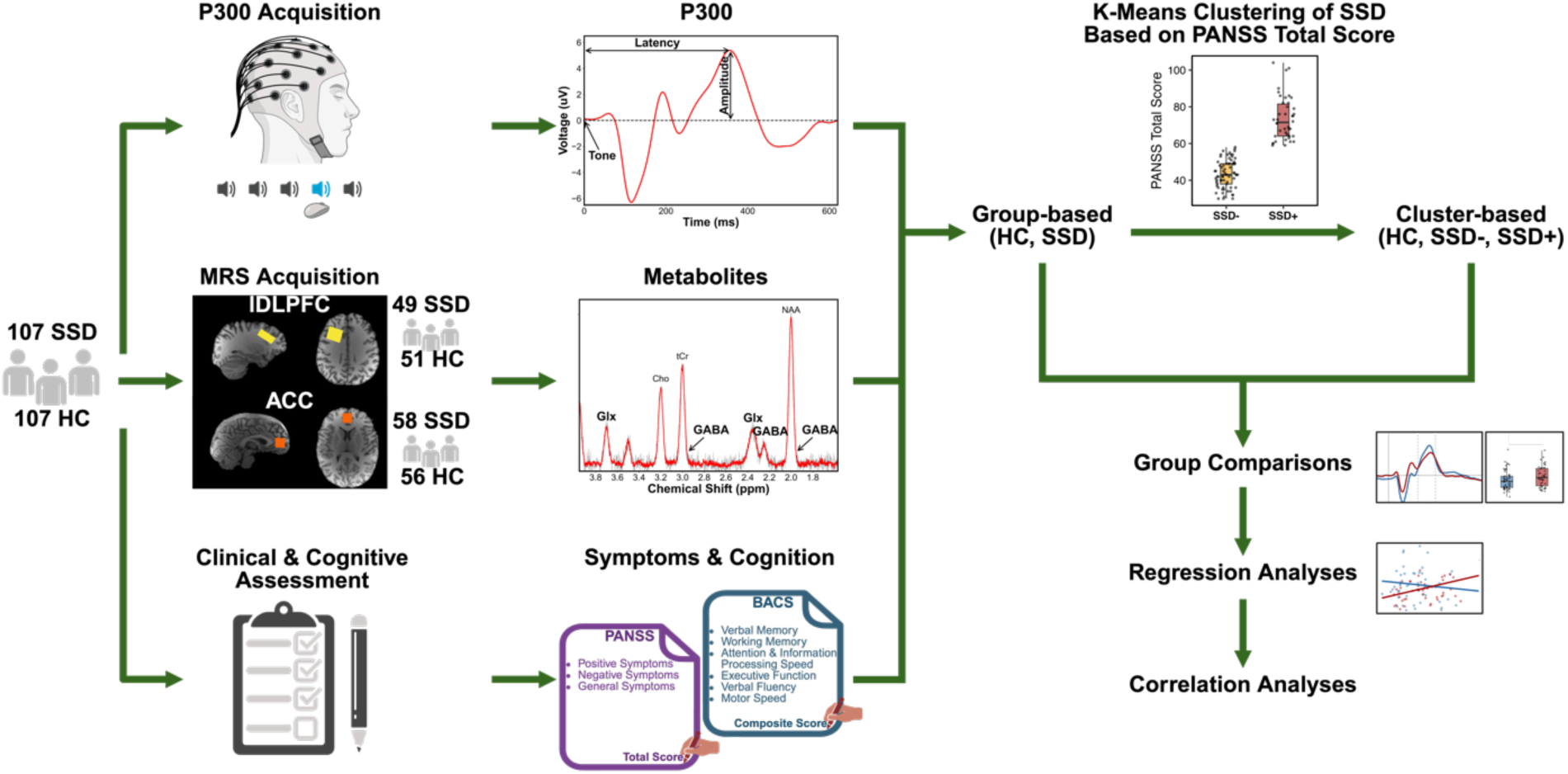
A summary diagram of the data acquisition and analysis. HC = healthy controls, SSD = schizophrenia spectrum disorder, SSD+ = higher-symptom cluster, SSD- = lower-symptom cluster, ACC = anterior cingulate cortex, lDLPFC = left dorsolateral prefrontal cortex, P300 = positive peak around 300 ms during the auditory oddball task, GABA = gamma-aminobutyric acid, Glx = glutamate-glutamine peak, BACS = Brief Assessment of Cognition in Schizophrenia, PANSS = The Positive and Negative Syndrome Scale.

## Results

### Clustering by PANSS Scores

To address heterogeneity within our SSD sample, we clustered patients into two subgroups based on PANSS total scores, resulting in three clusters for further analysis: HC, SSD− (lower-symptom, N = 70), and SSD+ (higher-symptom, N = 42). We included both group-based (HC and SSD) and cluster-based analyses (HC, SSD-, and SSD+) for all the analyses. The SSD− cluster had significantly lower PANSS positive (Contrast EMM = −6.63, 95% CI [−7.81, −5.44], *d* = 2.23, *p* < .001), PANSS negative (Contrast EMM = −8.70, 95% CI [−9.79, −7.61], *d* = 3.18, *p* < .001), and PANSS total scores (Contrast EMM = −29.69, 95% CI [−32.43, −26.95], *d* = 4.31, *p* < .001), reflecting fewer symptoms. Additionally, the SSD− cluster exhibited higher BACS composite z-scores (Contrast EMM = 0.90, 95% CI [0.38, 1.42], *d* = 0.71, *p* < .001), indicating better cognitive function. In summary, the SSD− cluster presented with fewer symptoms and better cognitive function compared to the SSD+ cluster, as indicated by lower PANSS scores and higher BACS composite z-scores.

### Behavioral and ERP Measures for P300

The SSD group demonstrated slower reaction times (RTs) than the HC group (*F*(1, 210) = 16.75, *p* < .001; mean HC = 339 ms, mean SSD = 386 ms) and a lower percentage of correct responses (*F*(1, 210) = 7.47, *p* = .007; mean HC = 98.6%, mean SSD = 95.7%) for the auditory oddball task.

Consistent with these behavioral impairments, the SSD group also exhibited decreased P300 amplitudes at central (*F*(1, 204) = 11.47, *p* < .001) and parietal (*F*(1, 208) = 32.27, *p* < .001) electrode sites (see Figure 2A). While there was a trend toward decreased frontal P300 amplitude in the SSD group, it did not reach statistical significance (*F*(1, 195) = 3.34, *p* = .069). No significant differences were observed in P300 latency across all regions (all *p*s > .05).

**Figure 2.**
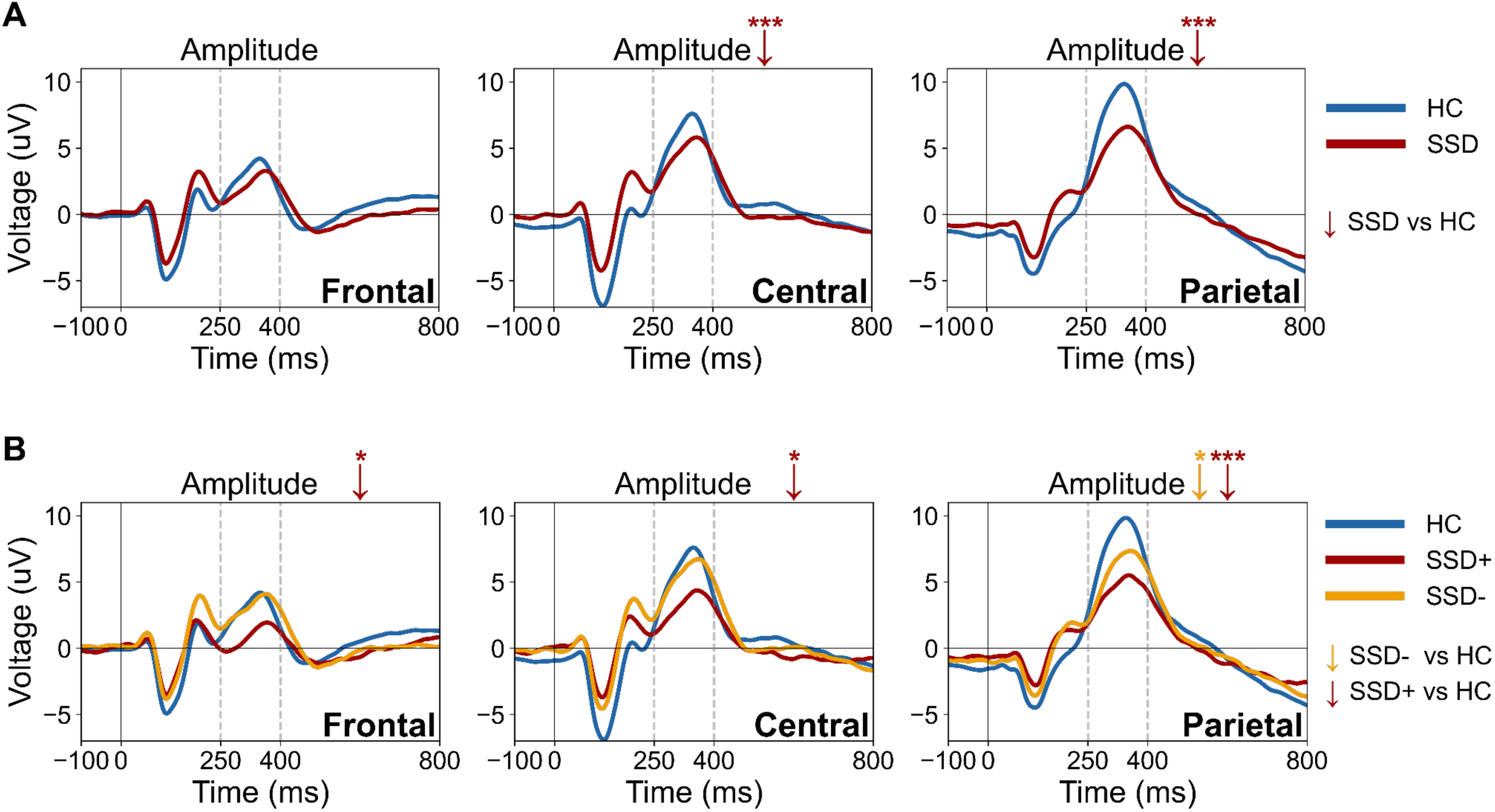
Frontal, central, and parietal ERPs showing P300 amplitude and latency differences across: **(A)** HC and SSD groups, and **(B)** HC, SSD-, and SSD+ clusters. The red (SSD, SSD+) and yellow (SSD-) arrows (↓) show the significant decrease in amplitude compared to HC. HC = healthy controls, SSD = schizophrenia spectrum disorder, SSD- = lower-symptom cluster, SSD+ = higher-symptom cluster, ms = milliseconds, μV = microvolts. Significance level: * < .05, ** < .01, *** < .001.

P300 amplitude comparisons of clusters further highlighted distinct electrophysiological profiles among the clusters. Compared to the HC cluster, the SSD+ cluster had significantly lower P300 amplitudes across all regions: frontal (Contrast EMM = −1.55, 95% CI [−3.06, −0.03], *d* = 0.40, *p* = .046), central (Contrast EMM = −2.24, 95% CI [−4.12, −0.36], *d* = 0.45 *p* = .020), and parietal (Contrast EMM = −3.08, 95% CI [−4.83, −1.33], *d* = 0.66, *p* < .001) (see Figure 2B). In contrast, the SSD− cluster showed significantly lower P300 amplitudes compared to HC only in the parietal region (Contrast EMM = −1.70, 95% CI [−3.20, −0.19], *d* = 0.36, *p* = .027) (see Figure 2B). No significant differences were observed in latencies for any comparison. Additionally, there were no significant behavioral differences during the task between the SSD− and SSD+ clusters in terms of RTs or percentage of correct responses (all *p*s > .05).

### Metabolite Comparisons

No significant group effects were observed for GABA or Glx in the ACC, nor for Glx in the lDLPFC (all *p*s > .05) when comparing HC and overall SSD, although there was a trend toward higher lDLPFC GABA levels in patients, it did not reach statistical significance (*F*(1, 94) = 3.65, *p* = .059; mean HC = 1.25, mean SSD = 1.39) (see Figure 3A).

**Figure 3.**
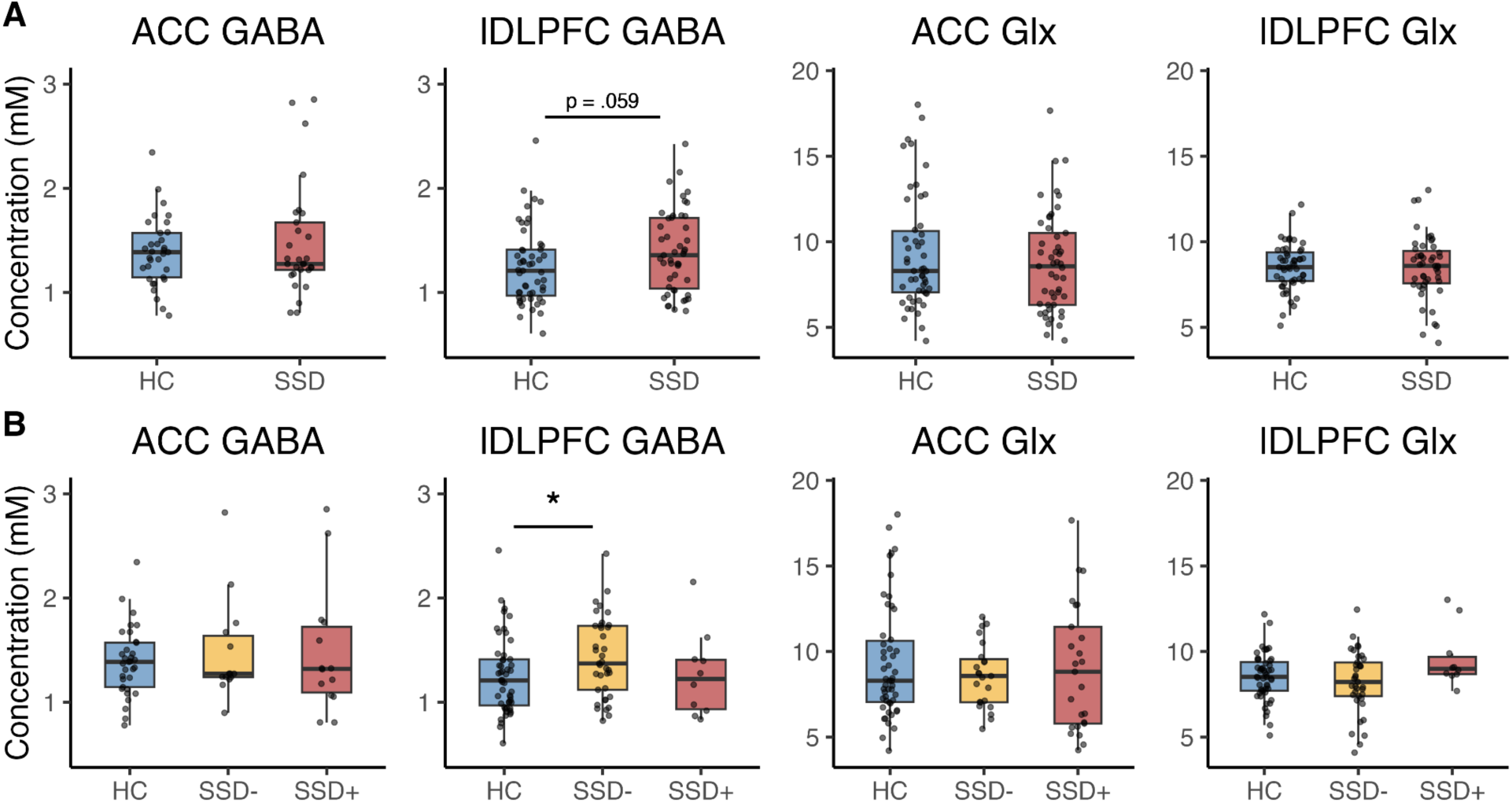
ACC and lDLPFC GABA and Glx levels across: **(A)** HC and SSD groups, and **(B)** HC, SSD-, and SSD+ clusters. HC = healthy controls, SSD = schizophrenia spectrum disorder, SSD- = lower-symptom cluster, SSD+ = higher-symptom cluster, ACC = anterior cingulate cortex, lDLPFC = left dorsolateral prefrontal cortex, GABA = gamma-aminobutyric acid, Glx = glutamate-glutamine peak, mM = millimolar. Significance level: * < .05, ** < .01, *** < .001.

An ANCOVA with the three clusters (HC, SSD−, SSD+) indicated a significant cluster effect for lDLPFC Glx levels (*F*(2, 94) = 3.42, *p* = .037) and a trend-level cluster effect for lDLPFC GABA levels (*F*(2, 93) = 2.63, *p* = .078). Due to the small sample size of SSD+ subjects with lDLPFC metabolite data (N = 10), which could introduce bias from unequal cluster sizes^36^, we conducted an additional ANCOVA including only the HC and SSD− clusters. This follow-up analysis revealed a significant increase in lDLPFC GABA levels in the SSD− cluster compared to HC (*F*(1, 84) = 5.15, *p* = .026), while lDLPFC Glx levels did not significantly differ between the SSD− and HC clusters (*F*(1, 85) = 1.50, *p* = .225) (see Figure 3B). No significant metabolite differences were observed in the ACC across clusters.

### Group-Based (overall SSD and HC) Regression and Correlation Analyses

To explore whether metabolite levels could account for the P300 amplitude differences observed between groups at central and parietal electrodes, we conducted linear regression analyses. The interaction between GABA levels in the lDLPFC and group was significantly associated with central P300 amplitude (B = 6.11, 95% CI [1.03, 11.11], *p* = .018), with the association being significant only for the SSD group, where increased GABA levels were associated with increased amplitude (B = 4.44, 95% CI [1.72, 7.19], *p* = .002) (see Figure 4A). A similar pattern was observed for parietal P300 amplitude, where the interaction between GABA levels in the lDLPFC and group was associated with amplitude (B = 4.78, 95% CI [0.09, 9.45], *p* = .045), with significance again limited to the SSD group, and increased GABA levels were associated with increased amplitude (B = 3.16, 95% CI [0.52, 5.81], *p* = .020) (see Figure 4A). However, GABA levels in the ACC were not associated with any P300 amplitudes, and Glx levels in either the lDLPFC or ACC did not demonstrate any significant relationship with P300 amplitudes.

**Figure 4.**
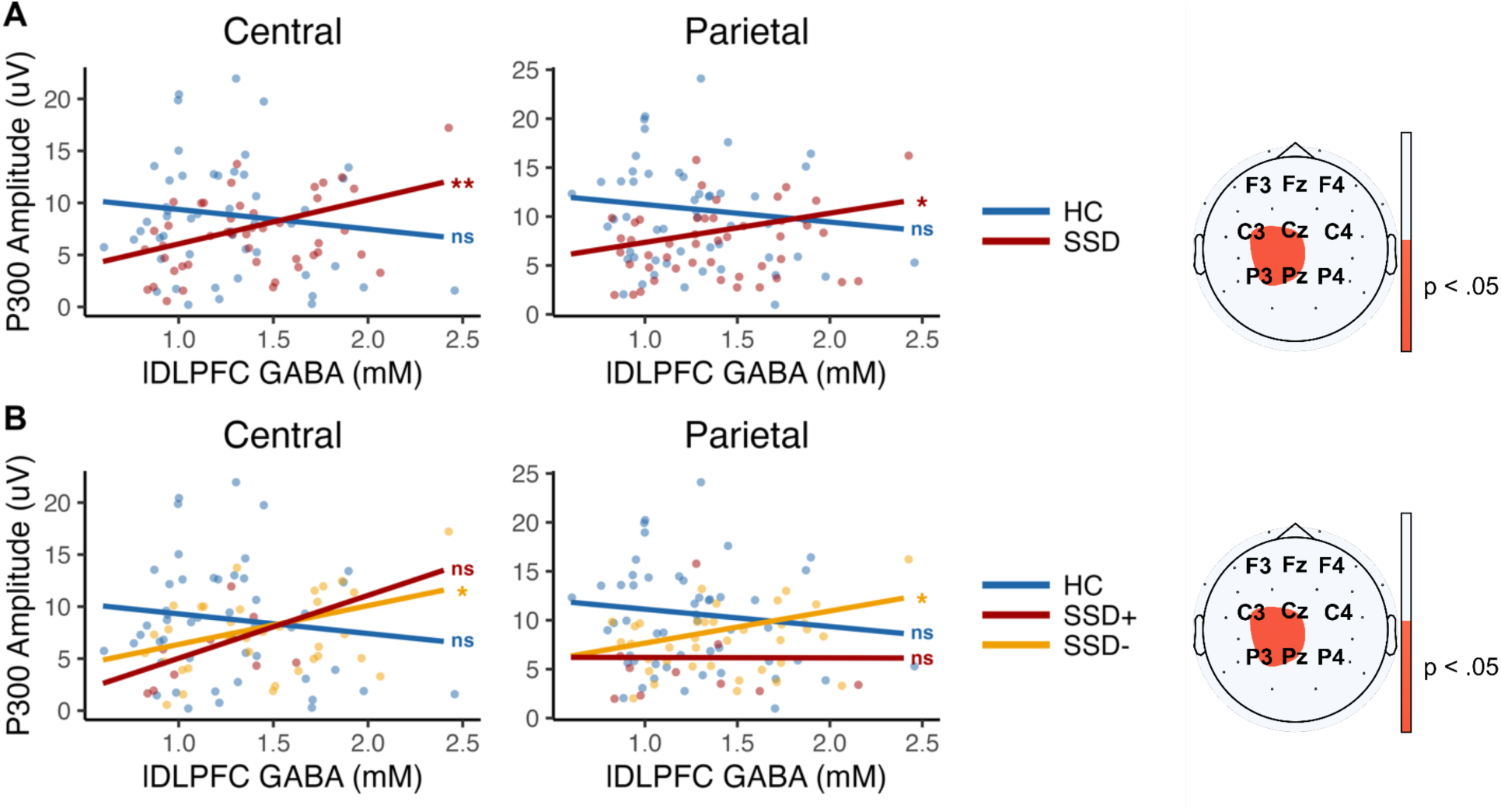
Central and parietal P300 amplitude and GABA relationship on the left, and significant electrodes for these relationships on the right across: **(A)** HC and SSD groups, and **(B)** HC, SSD-, and SSD+ clusters. HC = healthy controls, SSD = schizophrenia spectrum disorder, SSD- = lower-symptom cluster, SSD+ = higher-symptom cluster, μV = microvolts, mM = millimolar. Central electrodes: C3, Cz, C4. Parietal electrodes: P3, Pz, P4. Electrodes are placed according to the international 10-20 system. Significance level: * < .05, ** < .01, *** < .001, ns = not significant.

Since our DLPFC ROI was in the left hemisphere, we analyzed i) C3, P3, ii) Cz, Pz and iii) C4, P4 electrodes separately for hemisphere-specific associations. The interaction between lDLPFC GABA levels and group was significantly associated with amplitudes for left (C3, P3) and midline (Cz, Pz) electrodes but not for right (C4, P4) electrodes, with all significant associations limited to the SSD group (see the Supplemental Results for detailed results).

To link these regional associations to behavior and cognition in the SSD group, we correlated lDLPFC GABA levels and P300 amplitudes at central and parietal sites with RTs, percentage of correct responses, and the BACS composite z-scores. Central and parietal P300 amplitudes were positively correlated with percentage of correct responses (central: *r* = 0.29, 95% CI [0.11, 0.46], *p* = .002; parietal: *r* = 0.36, 95% CI [0.18, 0.52], *p* < .001), while only parietal P300 amplitude showed a positive correlation with the BACS composite z-score (*r* = 0.22, 95% CI [0.02, 0.40], *p* = .034) and a negative correlation with RTs (*r* = −0.27, 95% CI [−0.44, −0.09], *p* = .005). Notably, GABA levels were not correlated with any of the behavioral or cognitive measures.

### Cluster-Based (SSD- and SSD+) Regression and Correlation Analyses

The interaction between GABA levels in the lDLPFC and cluster was significantly associated with central P300 amplitude (B = 5.62, 95% CI [0.40, 10.87], *p* = .036), with the effect being significant only in the SSD− cluster, where increased GABA levels were associated with increased amplitude (B = 3.73, 95% CI [0.51, 6.95], *p* = .021) (see Figure 4B). Additionally, the interaction between lDLPFC GABA levels and cluster was significantly associated with parietal P300 amplitude (B = 5.06, 95% CI [0.04, 10.01], *p* = .048) (see Figure 4B). When modeled separately for each cluster, a significant association was observed only in the SSD− cluster, with increased GABA levels associated with increased amplitude (B = 3.16, 95% CI [0.32, 6.08], *p* = .030). Consistent with prior analyses, no significant relationships were found in models involving GABA levels in the ACC or Glx levels in the lDLPFC or ACC.

Similar to our group-based analysis, we analyzed i) C3, P3, ii) Cz, Pz and iii) C4, P4, electrodes separately to explore potential hemisphere-specific associations. The interaction between lDLPFC GABA levels and cluster was significantly associated with amplitudes for left (C3, P3) and midline (Cz, Pz) electrodes but not for right (C4, P4) electrodes, with all significant associations limited to the SSD− cluster (see the Supplemental Results for detailed results).

To link these associations to behavior and cognition in the SSD− cluster, we performed correlation analyses between GABA levels in the lDLPFC and P300 amplitudes at central and parietal sites with RTs, percentage of correct responses, and the BACS composite z-scores. Central and parietal P300 amplitudes were positively correlated with percentage of correct responses (central: *r* = 0.30, 95% CI [0.06, 0.50], *p* = .018; parietal: *r* = 0.38, 95% CI [0.15, 0.57], *p* = .002), while only parietal P300 amplitude showed a negative correlation with RTs (*r* = −0.26, 95% CI [−0.47, −0.01], *p* = .043). No correlations were found between central P300 amplitudes and the BACS composite z-score. Similar to the SSD group, GABA levels were not correlated with any of the behavioral or cognitive measures in this cluster.

## Discussion

This study investigated the potential role of GABA and Glx levels in the ACC and lDLPFC in relation to P300 ERP differences in SSD, and their links to cognition and behavior. Clustering the SSD group into higher (SSD+) and lower (SSD-) symptom subgroups provides insight into the heterogeneous nature of GABAergic function and its impact on cognitive processes across different symptom profiles.

Analysis of P300 amplitudes revealed significant differences between the SSD and HC groups, with reduced amplitudes in the parietal and central regions in the SSD group. Subgroup analysis revealed that higher-symptom patients had lower amplitudes across all regions, while lower-symptom patients, with fewer symptoms and better cognitive function, exhibited reductions only in the parietal region. Regarding metabolites, lower-symptom patients had higher GABA levels in the lDLPFC compared to HC, whereas no significant differences were found in the overall SSD group. Higher GABA levels in the lDLPFC were associated with increased P300 amplitudes at central and parietal sites for the overall SSD and lower-symptom groups. Although GABA levels were linked to P300 amplitudes, which in turn positively correlated with the BACS composite score and behavioral performance, GABA itself did not directly correlate with behavioral or cognitive measures indicating rather indirect impact on the neurophysiology and cognition outcome in this cohort.

### Amplitude Differences Across Regions

P300 amplitude comparisons revealed distinct ERP profiles between patients and healthy controls, with higher and lower-symptom patients showing different patterns of impairment. P300 amplitude was significantly reduced in the SSD group compared to HC in the parietal and central regions, aligning with the well-replicated amplitude reductions in SSD^3,4^. Higher-symptom patients exhibited significantly lower P300 amplitudes than healthy controls across all regions (frontal, central, and parietal), suggesting widespread neurophysiological impairment. In contrast, lower-symptom patients had reduced amplitudes compared to controls only in the parietal region, with preserved amplitudes in frontal and central areas. This pattern aligns with the literature showing that the degree of P300 reduction varies with symptom severity^8,9^. Correlations between P300 amplitude and behavioral and cognitive performance linked higher amplitudes to better cognitive performance, consistent with previous research demonstrating the relationship between P300 and various cognitive domains (see Hamilton et al.^5^ for a review). Additionally, lower-symptom patients had higher BACS composite scores than higher-symptom patients, further highlighting the relationship between cognitive impairment and symptom severity. While P300 amplitudes correlated with cognitive measures, GABA levels showed no direct associations, suggesting that P300 may be a more immediate marker of cognitive function, with GABA’s influence potentially being indirect or mediated by other factors.

### GABAergic Dysfunction in the ACC and lDLPFC

We observed increased GABA levels in the lDLPFC specifically for lower-symptom patients but not for the broader SSD patient group. Kegeles et al.^37^ found no changes in lDLPFC GABA levels compared to controls, potentially due to differences in sample characteristics, including symptom severity and group sizes (PANSS total scores: 57 for medication-receiving [N = 7], 71 for medication-free [N = 15]), compared to our lower-symptom group with lDLPFC GABA measurement (PANSS total scores: 42 [N = 38]). On the other hand, our finding of altered GABA in the DLPFC aligns with postmortem studies showing alterations in GABA_A_ receptor binding, GAD67, GAD65, and GAT-1, which are involved in GABAergic transmission (see De Jonge et al.^38^ for a review).

Furthermore, we found a positive association between lDLPFC GABA levels and P300 amplitude within the patient group, particularly among the lower-symptom subgroup. While no previous studies have directly linked MRS-measured GABA to P300, our findings differ from studies on GABA agonists in healthy individuals, showing a negative GABA and P300 amplitude association^39–41^. This distinction may highlight the differences between acute GABA modulation in healthy subjects and chronic GABA alterations observed in SSD. Unlike acute GABA increases in healthy subjects, which can temporarily suppress cortical activity, chronically elevated GABA in SSD may reflect a compensatory mechanism aimed at restoring the E/I balance. Additionally, the observed positive relationship in our patients, absent in controls, highlights how GABA-related modulation of P300 may differ fundamentally between SSD and HC, possibly reflecting the distinct neuroadaptive processes in SSD.

Our findings support prior studies highlighting the DLPFC’s role in SSD pathophysiology, particularly in cognitive domains such as working memory and cognitive control^42^. The DLPFC’s involvement in these domains makes it particularly sensitive to shifts in the excitatory-inhibitory (E/I) balance^43^, with a potential GABAergic compensation mechanism buffering against these imbalances. The E/I balance reflects the interaction between excitatory glutamatergic neurons and inhibitory GABAergic interneurons. Disruptions to this balance have been proposed as underlying mechanisms in neuropsychiatric disorders, including schizophrenia^10^. Previous studies suggest that GABAergic dysfunction affects the DLPFC’s capacity to regulate excitatory inputs, thereby supporting cognitive functions^44^. Increased GABA levels in our lower-symptom patients could represent an adaptive mechanism to stabilize dysfunction and maintain cognitive control, as suggested by more preserved P300 amplitudes and cognitive performance in this group. In contrast, the higher-symptom group may lack sufficient GABAergic compensation, leading to broader cognitive deficits, where increased inhibition alone no longer stabilizes the E/I balance.

In contrast to the lDLPFC, ACC GABA levels did not differ from healthy controls in patients or subgroups. Prior MRS studies have shown increased GABA levels in the ACC for ultra-high-risk (UHR) individuals, patients with first-episode psychosis (FEP), and medication-naïve patients with schizophrenia compared to healthy controls^37,45–48^, highlighting the role of GABAergic dysfunction or compensation in early stages of illness or in medication-naïve conditions. However, our sample consisted of mostly chronic patients receiving antipsychotic treatment. A study with a chronic schizophrenia sample reported increased ACC GABA/Cr ratios; however, creatine (Cr) as an internal reference has been questioned due to its potential fluctuations in disease states^49^. Furthermore, in the study of Kegeles et al.^37^, although ACC GABA was increased in the medication-free group, the medication-receiving group did not show any differences with healthy controls. Similarly, an intervention study on FEP patients found that while ACC GABA levels were elevated at baseline compared to healthy people, these differences disappeared after four weeks of antipsychotic treatment^50^ likely due to medication normalization. Together, these findings and study population differences suggest that antipsychotic treatment and longer duration of the illness may stabilize or normalize GABA levels in the ACC, potentially accounting for the lack of significant changes observed in our sample.

Overall, our findings suggest a potential region-specific compensatory mechanism in GABA functioning. This mechanism may optimize the excitation-inhibition balance, enhancing cognitive processing in less symptomatic patients. However, this compensatory mechanism may break down with illness progression or higher symptom severity, as seen in our higher-symptom group. While acute increases in GABAergic inhibition are likely to suppress cortical activity, in SSD, persistent region-specific GABA elevations might stabilize the E/I balance over time, supporting cognitive functioning.

### Glx Levels and the Role of Glutamatergic Dysfunction

Unlike GABA, Glx was not linked to P300 amplitude and showed no significant differences between SSD and HC in either the ACC or lDLPFC. Although findings on glutamatergic levels are mixed, most studies show that patients receiving antipsychotic medication have Glx levels comparable to healthy controls (see Poels et al.^51^ for a review). The absence of Glx group differences and Glx-P300 amplitude relationships in our study does not rule out the relevance of glutamatergic dysfunction and its link to cognition in SSD. MRS primarily captures intracellular glutamate levels and cannot directly assess synaptic transmission or excitatory signaling^52,53^. Bojesen et al.^54^ further suggest that cognitive deficits in schizophrenia may be linked to impaired glutamate dynamics during cognitive tasks rather than at resting levels. Additionally, NMDAR hypofunction has been implicated in cognitive deficits and P300 abnormalities in schizophrenia^5^. The role of glutamate-glutamine in task-specific contexts or in receptor-mediated processes highlights an important direction for further research on SSD-related cognitive deficits.

### Limitations, Strengths, and Future Directions

This study has several limitations. First, medication effects remain a concern, and although we did not find any relationships between GABA levels or P300 amplitudes and chlorpromazine equivalent dose (CPZeq, see the Supplemental Results), this measure does not capture differences in drug classes, and its longitudinal changes remain unknown in this study. Second, the smaller SSD+ group size limits statistical power, and while our overall sample size exceeds many MRS studies, it remains a constraint for detecting smaller effects. Third, the lack of simultaneous MRS-EEG recordings and the use of resting-state rather than task-based MRS limit the assessment of dynamic neurochemical-electrophysiological interactions. Lastly, the cross-sectional design limits causal interpretations, highlighting the need for longitudinal research. Despite these limitations, this study benefits from a larger sample than most prior MRS studies, the integration of MRS, EEG, and cognitive measures, and a symptom-based subgroup analysis. Future research should address these limitations by incorporating larger, balanced samples, longitudinal designs, and simultaneous task-based MRS-EEG to capture dynamic metabolite-electrophysiology interactions, while also accounting for different drug classes and their longitudinal variations.

## Conclusion

These findings suggest that region-specific GABAergic alterations in the lDLPFC may be linked to differences in cognitive function and neurophysiology in SSD. The observed links between GABA, P300, and cognitive performance emphasize the complex interplay of neurochemical and electrophysiological processes in SSD. The variation in results across symptom profiles emphasizes the importance of considering heterogeneity within SSD subgroups when interpreting neurochemical and cognitive findings. While increased GABA in the lDLPFC of lower-symptom patients may reflect an adaptive mechanism supporting cognitive function, the absence of similar findings in higher-symptom patients suggests a potential breakdown of this process with greater illness severity. However, this study was purely exploratory, and this suggested mechanism should be tested in a follow-up hypothesis-driven study while accounting for the limitations we reported. In contrast, no significant alterations in ACC GABA were observed, which may reflect stabilization effects of chronic illness or antipsychotic treatment. These findings highlight the importance of considering regional and symptom-based differences.

## Data Availability

All data produced in the present study are available upon reasonable request to the authors

## Acknowledgements

The MRS package was developed by Edward J. Auerbach and Małgorzata Marjańska and provided by the University of Minnesota under a C2P agreement. The procurement of the MRI scanner was supported by the Deutsche Forschungsgemeinschaft (DFG, German Research Foundation) grant for major research (DFG, INST 86/1739-1 FUGG). This research was supported by BMBF with the EraNet project GDNF UpReg (01EW2206) to PF, AS, VY and GH. The study is funded by the EU HORIZON-INFRA-2024-TECH-01-04 project DTRIP4H 101188432 to PF, AS and FR. VY was supported by the Residency/PhD track of the International Max Planck Research School for Translational Psychiatry (IMPRS-TP). VY is supported by the Faculty of Medicine at LMU Munich (FöFoLe Reg.-Nr. 1226/2024). JM was supported by the Faculty of Medicine at LMU Munich (FöFoLe Reg.-Nr. 1167). The study was endorsed by the Federal Ministry of Education and Research (Bundesministerium für Bildung und Forschung [BMBF]) within the initial phase of the German Center for Mental Health (DZPG) (grant: 01EE2303A, 01EE2303F to PF). The study was funded by the Supplement to BMBF funding for the German Centre for Mental Health (DZPG) by the Bavarian State Ministry for Science and the Arts with the Grant for the research project Improving Infrastructures for DZPG and NAKO Cohorts to PF, DK and BK.

## Declaration of generative AI and AI-assisted technologies in th**e** writing process

During the preparation of this work the authors used the GPT – 4 model developed by OpenAI to improve readability and language of the manuscript. After using this tool/service, the authors reviewed and edited the content as needed and take full responsibility for the content of the published article.

## Author Contributions

D.K., E.W., and F.J.R. designed and conceptualized the Clinical Deep Phenotyping study. B.K., V.M., G.H., M.S.K., and D.K. formulated the idea and designed the current study. V.M., M.S.K., F.D., N.K., M.K., A.H., J.M., and S.S. recruited patients and collected study data. E.W., V.Y., and J.M. trained staff on diagnostic and clinical assessments. MRI measurements were performed by M.S.K., J.M., F.D., and S.S. under the supervision of D.K. and L.R. EEG measurements were performed by V.M., F.D., N.K., and M.K. under the supervision of D.K. B.K., V.M., M.S.K., and G.V. processed MRS data. B.K. and G.H. processed EEG data. Statistical analysis and visualization were done by B.K., V.M., and D.K. B.K., V.M., G.H., M.S.K., and D.K. wrote the manuscript. E.B., L.R., M.K., T.G., A.Š., G.V., V.Y., J.M., and F.J.R. provided critical review. B.K., V.M., and D.K. prepared the final manuscript with the help of all authors.

## CDP Working Group

Stephanie Behrens, Emanuel Boudriot, Man-Hsin Chang, Valéria de Almeida, Sylvia de Jonge, Fanny Dengl, Peter Falkai, Laura E. Fischer, Nadja Gabellini, Vanessa Gabriel, Sabrina Galinski, Thomas Geyer, Katharina Hanken, Alkomiet Hasan, Genc Hasanaj, Alexandra Hisch, Georgios Ioannou, Iris Jäger, Marcel S. Kallweit, Temmuz Karali, Susanne Karch, Berkhan Karslı, Daniel Keeser, Christoph Kern, Nicole L. Klimas, Maxim Korman, Nikolaos Koutsouleris, Lenka Krcmar, Verena Meisinger, Julian Melcher, Matin Mortazavi, Joanna Moussiopoulou, Karin Neumeier, Frank Padberg, Boris Papazov, Irina Papazova, Sergi Papiol, Pauline Pingen, Oliver Pogarell, Siegfried G. Priglinger, Florian J. Raabe, Lukas Roell, Moritz J. Rossner, Philipp Sämann, Andrea Schmitt, Susanne Schmölz, Eva C. Schulte, Enrico Schulz, Benedikt Schworm, Elias Wagner, Sven Wichert, Vladislav Yakimov, Peter Zill, Zhuanghua Shi, Michael J. Ziller

## Supplemental information

Document S1. Figures S1–S2, Tables S1-S2, Supplemental Methods (Clinical and Cognitive assessments; P300 Recording and Preprocessing), Supplemental Results (Midline Electrode P300 Comparisons; Hemisphere-Specific Regression Analyses; Smoking and CPZeq Effects on lDLPFC GABA Levels and P300 Amplitudes; Group Differences in Gray Matter Volume of the ACC and lDLPFC)

## Supplemental Methods

### Clinical and Cognitive assessments

Both patients and healthy controls underwent comprehensive clinical assessments. Basic socio-demographic data, family and psychiatric history, suicidality, and lifetime substance use were collected using the Munich Mental Health Biobank self-report questionnaire. Patient diagnoses were confirmed using the MINI International Neuropsychiatric Interview (v7.0.2) based on DSM-V criteria, and symptom severity was assessed with the Positive and Negative Syndrome Scale (PANSS)^1^. Additional information on psychiatric history, demographics including sex assigned at birth, lifestyle factors, somatic conditions, body mass index (BMI), and medications was obtained through self-reports and medical records. The Global Assessment of Functioning (GAF) scale assessed participants’ general functioning. The Andreasen criteria were used to differentiate remission status^2^. Antipsychotic doses were converted to chlorpromazine equivalents (CPZeq) using the defined daily dose method^3^. Smoking status was assessed using the Fagerström Test for Nicotine Dependence^4^.

Neurocognitive function was assessed using the Brief Assessment of Cognition in Schizophrenia (BACS), a validated tool for evaluating cognitive deficits in individuals with schizophrenia^5^. The BACS, administered exclusively to participants with native-level German proficiency, consisted of seven tasks measuring six cognitive domains: verbal memory (list learning), working memory (digit sequencing), motor speed (token motor task), verbal fluency (category instances and controlled oral word association), attention and information processing speed (symbol coding), and executive function (Tower of London test)^6^. The assessment took approximately 40 minutes to complete. For data analysis, z-scores were calculated for each task using the mean and standard deviation of the HC group. These task z-scores were averaged into a single score, which was then transformed into the BACS composite score with a final z-score transformation based on the HC mean and standard deviation of the average score^5^.

### P300 Recording and Preprocessing

Participants performed an auditory oddball task wearing Phillips headphones, pressing a button in response to infrequent target tones (2000 Hz, 85 dB, 20% of 700 trials) randomly placed among standard tones (1000 Hz, 80 dB, 80% of trials) while keeping their eyes closed for 18 minutes. Tones were generated via Presentation software (neurobs.com; v14.9), lasting 40 ms with a 10-ms rise and fall time, and presented with a 1.5-second interstimulus interval. Data were converted to BIDS format using BV2BIDS (Brain Products, Martinsried, Germany).

EEG signals were re-referenced to the left and right mastoids, downsampled to 256 Hz, bandpass filtered between 0.05 and 70 Hz, and notch filtered at 49.5–50.5 Hz. The data were then segmented into 1.5-second epochs (−500 ms to 1000 ms relative to stimulus onset). Artifact removal involved a multi-step approach: epochs exceeding ±5 SD, showing linear trends (max slope=5, min R2=0.7), or containing low (<2 Hz; power threshold −50 to 50 dB) or high (20–40 Hz; power threshold −100 to 25 dB) frequencies were excluded. Channels were evaluated for poor quality using power spectrum (−4 to 6 std), kurtosis (−7 to 15 std), and joint probability (−9 to 7 std) thresholds. When >50% of epochs were marked for rejection, priority was given to channel removal, followed by epoch rejection based on the initial data. Datasets were excluded if >50% of data or >20% of electrodes were rejected. Independent component analysis (ICA) was then performed, and artifact components were removed using MARA^7^. Finally, a second round of rejection with adjusted thresholds (power spectrum [−6 to 5 std], kurtosis [−6 to 9 std], and joint probability [−7 to 7 std]) was conducted, and interpolations were applied to the removed channels. Baseline correction was performed using a −100 to 0 ms prestimulus window, and epochs exceeding ±100 µV were excluded. Cleaned data were converted back to continuous format for further analysis.

## Supplemental Results

### Midline Electrode P300 Comparisons

The SSD group exhibited decreased P300 amplitudes across all three midline electrodes: Fz (*F*(1, 195) = 5.55, *p* = .019), Cz (*F*(1, 200) = 9.36, *p* = .003), and Pz (*F*(1, 208) = 28.88, *p* < .001) (see Figure S1A). No significant differences were observed in P300 latency across midline electrodes (all *p*s > .05).

P300 amplitude comparisons of clusters further highlighted distinct electrophysiological profiles among the clusters. Compared to the HC cluster, the SSD+ cluster had significantly lower P300 amplitudes across all midline electrodes: Fz (Contrast EMM = −2.20, 95% CI [−4.20, −0.21], *d* = 0.43, *p* = .030), Cz (Contrast EMM = −2.27, 95% CI [−4.48, −0.06], *d* = 0.40, *p* = .045), and Pz (Contrast EMM = −3.15, 95% CI [−5.09, −1.21], *d* = 0.61, *p* = .002) (see Figure S1B). In contrast, the SSD− cluster showed significantly lower P300 amplitudes compared to HC only in the Pz electrode (Contrast EMM = −1.81, 95% CI [−3.48, −0.14], *d* = 0.35, *p* = .034) (see Figure S1B). No significant differences were observed in latencies for any comparison.

### Hemisphere-Specific Regression Analyses

To investigate hemisphere-specific associations given our DLPFC ROI was in the left hemisphere, we analyzed C3, Cz, C4, P3, Pz, and P4 electrodes separately. Significant interactions between GABA levels in the lDLPFC and group were observed for left (C3, P3) and midline (Cz, Pz) electrodes in the central and parietal regions, but not for right electrodes. The interaction between GABA levels in the lDLPFC and group significantly predicted amplitude for C3 (B = 5.78, 95% CI [1.09, 10.42], *p* = .017), Cz (B = 6.98, 95% CI [1.07, 12.82], *p* = .021), P3 (B = 5.22, 95% CI [0.69, 9.77], *p* = .024), and Pz (B = 5.63, 95% CI [0.30, 11.04], *p* = .036). The GABA levels in the lDLPFC significantly predicted amplitudes only in the SSD group (C3: B = 3.46, 95% CI [0.69, 6.31], *p* = .014; Cz: B = 5.14, 95% CI [1.74, 8.48], *p* = .003; P3: B = 3.50, 95% CI [0.88, 6.17], *p* = .007; Pz: B = 3.42, 95% CI [0.38, 6.45], *p* = .027). No significant interaction associations were observed for right hemisphere electrodes of central and parietal regions (C4, P4; ps > .05).

We repeated the same analysis for cluster-based groups. As in the previous analysis, significant associations were observed only for left (C3, P3) and midline (Cz, Pz) electrodes, but not for right electrodes. The interaction between lDLPFC GABA levels and cluster significantly predicted amplitudes for C3 (B = 6.34, 95% CI [1.51, 11.35], *p* = .011), Cz (B = 6.40, 95% CI [0.15, 12.64], *p* = .046), P3 (B = 5.61, 95% CI [0.81, 10.32], *p* = .024), and Pz (B = 6.02, 95% CI [0.43, 11.63], *p* = .032). These associations were significant only in the SSD− cluster (C3: B = 3.90, 95% CI [0.60, 7.18], *p* = .022; Cz: B = 4.32, 95% CI [0.41, 8.33], *p* = .029; P3: B = 3.52, 95% CI [0.60, 6.50], *p* = .016; Pz: B = 3.44, 95% CI [0.24, 6.59], *p* = .032). No significant associations were observed for right hemisphere electrodes of the central and parietal regions (C4, P4; *p*s > .05).

### Smoking and CPZeq Effects on lDLPFC GABA Levels and P300 Amplitudes

We examined the correlation between CPZeq and lDLPFC GABA levels, as well as P300 amplitudes in the central and parietal regions, within the SSD group, including age and sex as covariates. No significant correlations were observed (*p*s > .05).

We ran an ANCOVA with age and sex as covariates to compare P300 amplitudes and lDLPFC GABA levels based on smoking status (smoker vs. non-smoker) within the SSD group. Smoking significantly decreased P300 amplitudes in all three regions: frontal (*F*(1, 94) = 10.69, *p* = .002), central (*F*(1, 100) = 9.64, *p* = .003), and parietal (*F*(1, 101) = 5.13, *p* = .026). However, lDLPFC GABA levels did not differ significantly based on smoking status (*p* > .05). We then included smoking status as a covariate, in addition to age and sex, and reran the group-based (overall SSD and HC) and cluster-based (SSD− and SSD+) regression analyses.

For the group-based regression, including smoking status as a covariate, the interaction between GABA levels in the lDLPFC and group was significantly associated with central P300 amplitude (B = 6.14, 95% CI [1.10, 11.04], *p* = .017), with the association being significant only in the SSD group, where increased GABA levels were associated with increased amplitude (B = 4.33, 95% CI [1.58, 7.11], *p* = .002). A similar pattern was observed for parietal P300 amplitude, where the interaction between GABA levels in the lDLPFC and group was significantly associated with amplitude (B = 4.74, 95% CI [−0.02, 9.56], *p* = .050). This association was again limited to the SSD group, where increased GABA levels were associated with increased amplitude (B = 3.14, 95% CI [0.55, 5.77], *p* = .021).

For the cluster-based regression, including smoking status as a covariate, the interaction between GABA levels in the lDLPFC and cluster was significantly associated with central P300 amplitude (B = 5.66, 95% CI [0.48, 10.90], *p* = .031), with the effect being significant only in the SSD− cluster, where increased GABA levels were associated with increased amplitude (B = 3.82, 95% CI [0.58, 7.03], *p* = .020). Additionally, the interaction between lDLPFC GABA levels and cluster was significantly associated with parietal P300 amplitude (B = 5.07, 95% CI [0.02, 10.08], *p* = .049). When modeled separately for each cluster, a significant association was observed only in the SSD− cluster, where increased GABA levels were associated with increased amplitude (B = 3.23, 95% CI [0.29, 6.12], *p* = .029).

We found no significant associations between CPZeq and lDLPFC GABA levels or P300 amplitudes. Smoking was significantly associated with reduced P300 amplitudes, but controlling for smoking status did not affect the main findings of either the group-based or cluster-based analyses.

### Group Differences in Gray Matter Volume of the ACC and lDLPFC

We examined gray matter volume (GMV) differences in the ACC and lDLPFC across groups in our cohort using ANCOVA, with age and sex included as covariates. We included the A46L, A946dL, A946vL, A8vlL, and A9lL regions for the lDLPFC and the A32pL and A32pR regions for the ACC, based on the Human Brainnetome Atlas^8^. No significant differences were observed in lDLPFC GMV (all *p*s > .05). However, ACC volumes were significantly decreased in the SSD group compared to HC (Contrast EMM = −110.30, 95% CI [−207.09, −13.51], *d* = 0.32, *p* = .026), with similar reductions observed in the SSD− (Contrast EMM = −116.60, 95% CI [−226.23, −6.96], *d* = 0.34, *p* = .037); but not in the SSD+(Contrast EMM = −115.97, 95% CI [−245.12, 13.18], *d* = 0.34, *p* = .078) clusters compared to HC (see Figure S2). No significant differences in ACC volumes were observed between the SSD− and SSD+clusters (*p* > .05).

## Supplemental Tables

**Table S1.**
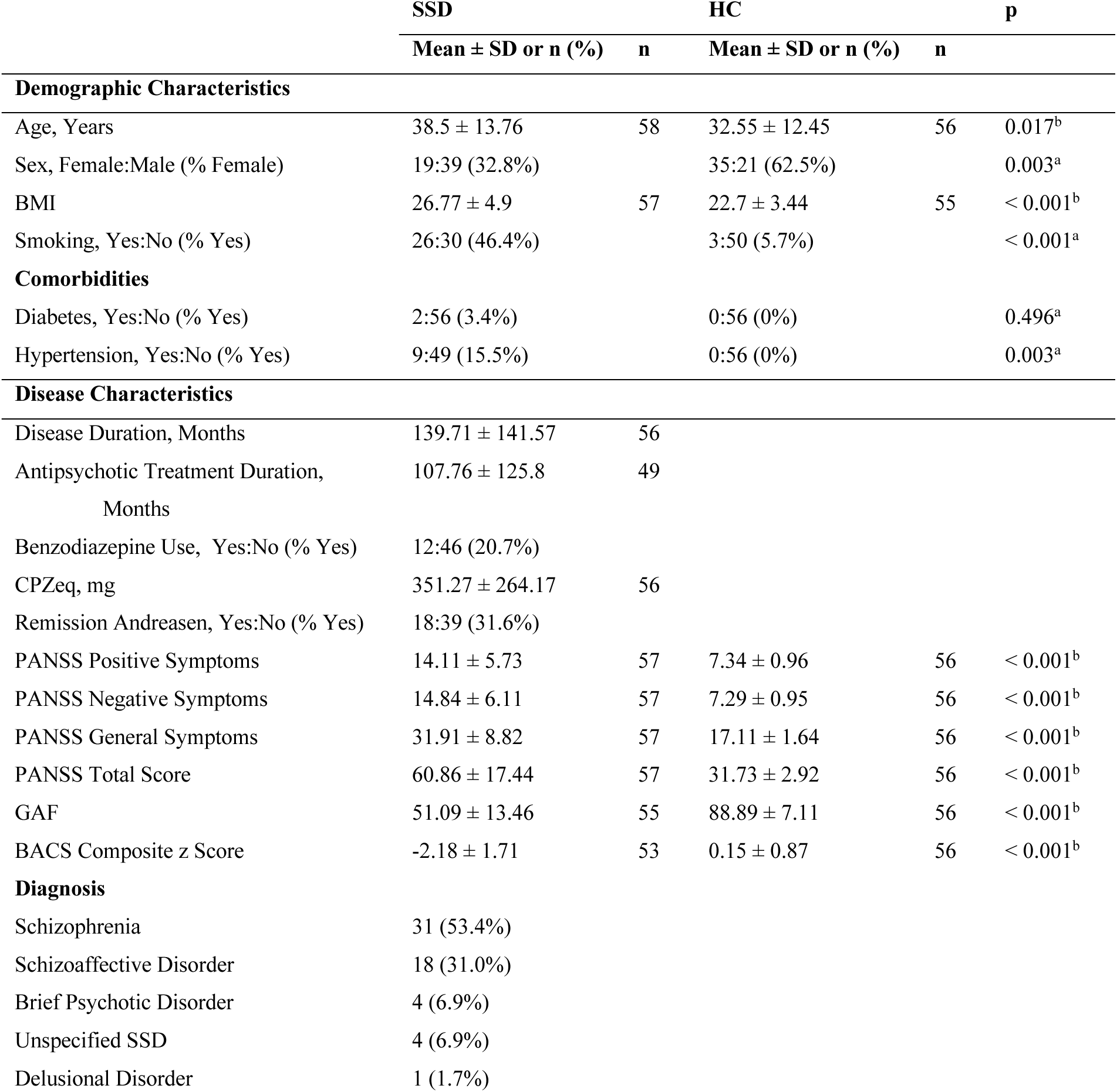
Sample characteristics table for the ACC MRS measurement. HC = healthy control participant, SSD = schizophrenia spectrum disorder, BACS = Brief Assessment of Cognition in Schizophrenia, BMI = body mass index, CPZeq = chlorpromazine equivalent dose, GAF = Global Assessment of Functioning, PANSS = Positive and Negative Syndrome Scale. *^a^* Fisher’s exact test, *^b^* Welch’s t test.

|  | SSD |  | HC |  | p |
| --- | --- | --- | --- | --- | --- |
| | Mean ± SD or n (%) | n | Mean ± SD or n (%) | n | |
| **Demographic Characteristics** |  |  |  |  |  |
| Age, Years | 38.5 ± 13.76 | 58 | 32.55 ± 12.45 | 56 | 0.017<sup>b</sup> |
| Sex, Female:Male (% Female) | 19:39 (32.8%) |  | 35:21 (62.5%) |  | 0.003<sup>a</sup> |
| BMI | 26.77 ± 4.9 | 57 | 22.7 ± 3.44 | 55 | < 0.001<sup>b</sup> |
| Smoking, Yes:No (% Yes) | 26:30 (46.4%) |  | 3:50 (5.7%) |  | < 0.001<sup>a</sup> |
| **Comorbidities** |  |  |  |  |  |
| Diabetes, Yes:No (% Yes) | 2:56 (3.4%) |  | 0:56 (0%) |  | 0.496<sup>a</sup> |
| Hypertension, Yes:No (% Yes) | 9:49 (15.5%) |  | 0:56 (0%) |  | 0.003<sup>a</sup> |
| **Disease Characteristics** |  |  |  |  |  |
| Disease Duration, Months | 139.71 ± 141.57 | 56 | | | |
| Antipsychotic Treatment Duration, Months | 107.76 ± 125.8 | 49 | | | |
| Benzodiazepine Use, Yes:No (% Yes) | 12:46 (20.7%) |  |  |  |  |
| CPZeq, mg | 351.27 ± 264.17 | 56 | | | |
| Remission Andreasen, Yes:No (% Yes) | 18:39 (31.6%) |  |  |  |  |
| PANSS Positive Symptoms | 14.11 ± 5.73 | 57 | 7.34 ± 0.96 | 56 | < 0.001<sup>b</sup> |
| PANSS Negative Symptoms | 14.84 ± 6.11 | 57 | 7.29 ± 0.95 | 56 | < 0.001<sup>b</sup> |
| PANSS General Symptoms | 31.91 ± 8.82 | 57 | 17.11 ± 1.64 | 56 | < 0.001<sup>b</sup> |
| PANSS Total Score | 60.86 ± 17.44 | 57 | 31.73 ± 2.92 | 56 | < 0.001<sup>b</sup> |
| GAF | 51.09 ± 13.46 | 55 | 88.89 ± 7.11 | 56 | < 0.001<sup>b</sup> |
| BACS Composite z Score | -2.18 ± 1.71 | 53 | 0.15 ± 0.87 | 56 | < 0.001<sup>b</sup> |
| **Diagnosis** |  |  |  |  |  |
| Schizophrenia | 31 (53.4%) |  |  |  |  |
| Schizoaffective Disorder | 18 (31.0%) |  |  |  |  |
| Brief Psychotic Disorder | 4 (6.9%) |  |  |  |  |
| Unspecified SSD | 4 (6.9%) |  |  |  |  |
| Delusional Disorder | 1 (1.7%) |  |  |  |  |

**Table S2.** Sample characteristics table for the lDLPFC MRS measurement. HC = healthy control participant, SSD = schizophrenia spectrum disorder, BACS = Brief Assessment of Cognition in Schizophrenia, BMI = body mass index, CPZeq = chlorpromazine equivalent dose, GAF = Global Assessment of Functioning, PANSS = Positive and Negative Syndrome Scale. *^a^* Fisher’s exact test, *^b^* Welch’s t test.

|  | SSD |  | HC |  | p |
| --- | --- | --- | --- | --- | --- |
|  | Mean ± SD or n (%) | n | Mean ± SD or n (%) | n |  |
| Demographic Characteristics |  |  |  |  |  |
| Age, Years | 38.86 ± 10.31 | 49 | 35.45 ± 11.88 | 51 | 0.129<sup>b</sup> |
| Sex, Female:Male (% Female) | 13:36 (26.5%) |  | 20:31 (39.2%) |  | 0.206<sup>a</sup> |
| BMI | 30.43 ± 5.91 | 48 | 24.21 ± 3.08 | 50 | < 0.001<sup>b</sup> |
| Smoking, Yes:No (% Yes) | 27:22 (55.1%) |  | 5:45 (10%) |  | < 0.001<sup>a</sup> |
| Comorbidities |  |  |  |  |  |
| Diabetes, Yes:No (% Yes) | 2:47 (4.1%) |  | 1:50 (2%) |  | 0.614<sup>a</sup> |
| Hypertension, Yes:No (% Yes) | 8:41 (16.3%) |  | 4:47 (7.8%) |  | 0.23<sup>a</sup> |
| Disease Characteristics |  |  |  |  |  |
| Disease Duration, Months | 145.96 ± 96.01 | 49 |  |  |  |
| Antipsychotic Treatment Duration, Months | 140.52 ± 95.47 | 46 |  |  |  |
| Benzodiazepine Use, Yes:No (% Yes) | 1:48 (2%) |  |  |  |  |
| CPZeq, mg | 333.83 ± 232.96 | 48 |  |  |  |
| Remission Andreasen, Yes:No (% Yes) | 33:16 (67.3%) |  |  |  |  |
| PANSS Positive Symptoms | 10.94 ± 4.15 | 49 | 7.18 ± 0.52 | 51 | < 0.001<sup>b</sup> |
| PANSS Negative Symptoms | 11.76 ± 4.73 | 49 | 7.57 ± 0.98 | 51 | < 0.001<sup>b</sup> |
| PANSS General Symptoms | 25.55 ± 7.76 | 49 | 17 ± 1.46 | 51 | < 0.001<sup>b</sup> |
| PANSS Total Score | 48.24 ± 14.58 | 49 | 31.75 ± 2.31 | 51 | < 0.001<sup>b</sup> |
| GAF | 56.04 ± 7.78 | 49 | 91 ± 5.59 | 51 | < 0.001<sup>b</sup> |
| BACS Composite z Score | -1.46 ± 1.44 | 46 | -0.14 ± 1.11 | 48 | < 0.001<sup>b</sup> |
| Diagnosis |  |  |  |  |  |
| Schizophrenia | 36 (73.5%) |  |  |  |  |
| Schizoaffective Disorder | 12 (24.5%) |  |  |  |  |
| Brief Psychotic Disorder | 1 (2.0%) |  |  |  |  |

## Supplemental Figures

**Figure S1.**
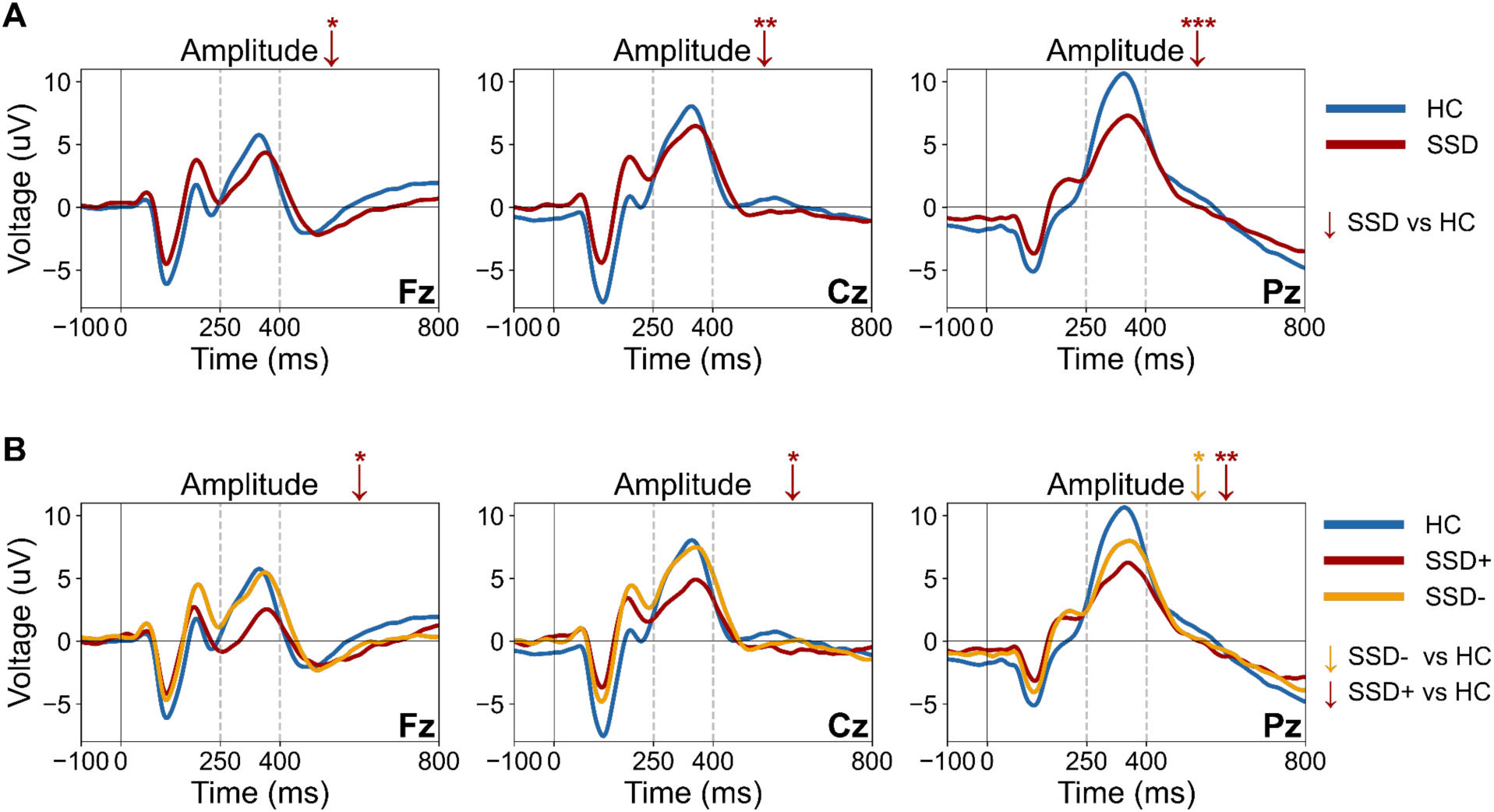
Fz, Cz, and Pz ERPs showing P300 amplitude and latency differences across: **(A)** HC and SSD groups, and **(B)** HC, SSD-, and SSD+ clusters. The red (SSD, SSD+) and yellow (SSD-) arrows (↓) show the significant decrease in amplitude compared to HC. HC = healthy controls, SSD = schizophrenia spectrum disorder, SSD- = lower-symptom cluster, SSD+ = higher-symptom cluster, ms = milliseconds, μV = microvolts. Significance level: * < .05, ** < .01, *** < .001.

**Figure S2.**
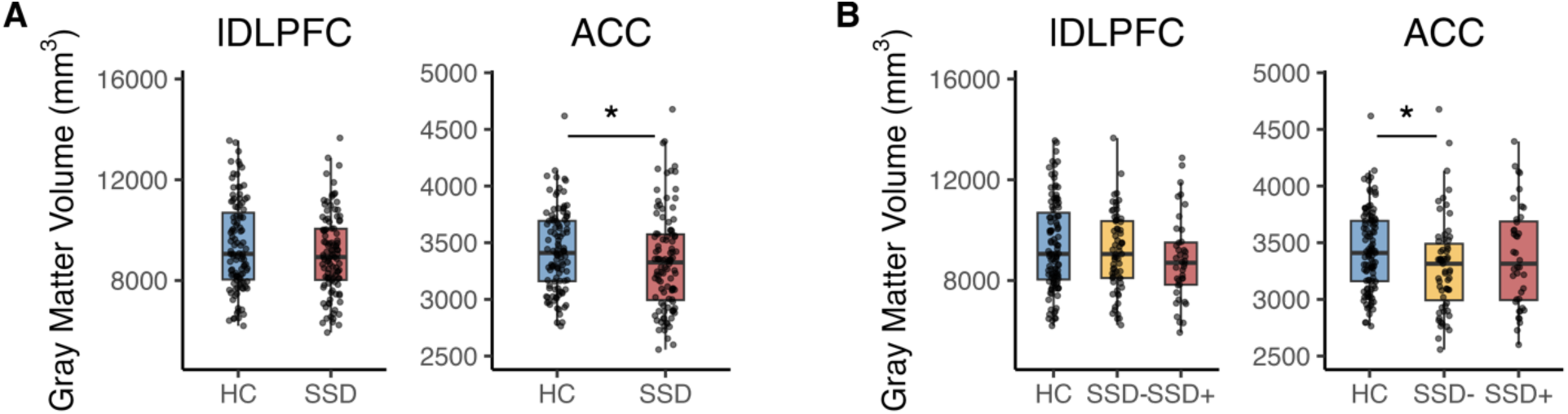
ACC and lDLPFC gray matter volumes across: **(A)** HC and SSD groups, and **(B)** HC, SSD-, and SSD+ clusters. HC = healthy controls, SSD = schizophrenia spectrum disorder, SSD- = lower-symptom cluster, SSD+ = higher-symptom cluster, ACC = anterior cingulate cortex, lDLPFC = left dorsolateral prefrontal cortex, mm^3^ = cubic millimeters. Significance level: * < .05, ** < .01, *** < .001.

## References

1. Kahn RS, Sommer IE, Murray RM, et al. Schizophrenia. Nat Rev Dis Primer. 2015;1(1):15067. doi:10.1038/nrdp.2015.67

2. McCutcheon RA, Keefe RSE, McGuire PK. Cognitive impairment in schizophrenia: aetiology, pathophysiology, and treatment. Mol Psychiatry. 2023;28(5):1902–1918. doi:10.1038/s41380-023-01949-9

3. Bramon E. Meta-analysis of the P300 and P50 waveforms in schizophrenia. Schizophr Res. 2004;70(2-3):315–329. doi:10.1016/j.schres.2004.01.004

4. Qiu Y qin, Tang Y xiang, Chan RCK, Sun X yang, He J. P300 Aberration in First-Episode Schizophrenia Patients: A Meta-Analysis. Chao L, ed. PLoS ONE. 2014;9(6):e97794. doi:10.1371/journal.pone.0097794

5. Hamilton HK, Mathalon DH, Ford JM. P300 in schizophrenia: Then and now. Biol Psychol. 2024;187:108757. doi:10.1016/j.biopsycho.2024.108757

6. Luck SJ. An Introduction to the Event-Related Potential Technique. 2nd ed. MIT Press; 2014.

7. De Lange FP, Heilbron M, Kok P. How Do Expectations Shape Perception? Trends Cogn Sci. 2018;22(9):764–779. doi:10.1016/j.tics.2018.06.002

8. Ford JM, Mathalon DH, Marsh L, et al. P300 amplitude is related to clinical state in severely and moderately ill patients with schizophrenia. Biol Psychiatry. 1999;46(1):94–101. doi:10.1016/S0006-3223(98)00290-X

9. Mathalon DH, Ford JM, Pfefferbaum A. Trait and state aspects of p300 amplitude reduction in schizophrenia: a retrospective longitudinal study. Biol Psychiatry. 2000;47(5):434–449. doi:10.1016/S0006-3223(99)00277-2

10. Liu Y, Ouyang P, Zheng Y, et al. A Selective Review of the Excitatory-Inhibitory Imbalance in Schizophrenia: Underlying Biology, Genetics, Microcircuits, and Symptoms. Front Cell Dev Biol. 2021;9:664535. doi:10.3389/fcell.2021.664535

11. Nakahara T, Tsugawa S, Noda Y, et al. Glutamatergic and GABAergic metabolite levels in schizophrenia-spectrum disorders: a meta-analysis of 1H-magnetic resonance spectroscopy studies. Mol Psychiatry. 2022;27(1):744–757. doi:10.1038/s41380-021-01297-6

12. Simmonite M, Steeby CJ, Taylor SF. Medial Frontal Cortex GABA Concentrations in Psychosis Spectrum and Mood Disorders: A Meta-analysis of Proton Magnetic Resonance Spectroscopy Studies. Biol Psychiatry. 2023;93(2):125–136. doi:10.1016/j.biopsych.2022.08.004

13. Picó-Pérez M, Vieira R, Fernández-Rodríguez M, De Barros MAP, Radua J, Morgado P. Multimodal meta-analysis of structural gray matter, neurocognitive and social cognitive fMRI findings in schizophrenia patients. Psychol Med. 2022;52(4):614–624. doi:10.1017/S0033291721005523

14. Zhao G, Lau WKW, Wang C, et al. A Comparative Multimodal Meta-analysis of Anisotropy and Volume Abnormalities in White Matter in People Suffering From Bipolar Disorder or Schizophrenia. Schizophr Bull. 2022;48(1):69–79. doi:10.1093/schbul/sbab093

15. Hertrich I, Dietrich S, Blum C, Ackermann H. The Role of the Dorsolateral Prefrontal Cortex for Speech and Language Processing. Front Hum Neurosci. 2021;15:645209. doi:10.3389/fnhum.2021.645209

16. Kelly AMC, Di Martino A, Uddin LQ, et al. Development of Anterior Cingulate Functional Connectivity from Late Childhood to Early Adulthood. Cereb Cortex. 2009;19(3):640–657. doi:10.1093/cercor/bhn117

17. Stevens FL, Hurley RA, Taber KH. Anterior Cingulate Cortex: Unique Role in Cognition and Emotion. Hurley RA, Hayman LA, Taber KH, eds. J Neuropsychiatry Clin Neurosci. 2011;23(2):121–125. doi:10.1176/jnp.23.2.jnp121

18. Minzenberg MJ, Laird AR, Thelen S, Carter CS, Glahn DC. Meta-analysis of 41 Functional Neuroimaging Studies of Executive Function in Schizophrenia. Arch Gen Psychiatry. 2009;66(8):811. doi:10.1001/archgenpsychiatry.2009.91

19. Li F, Wang J, Jiang Y, et al. Top-Down Disconnectivity in Schizophrenia During P300 Tasks. Front Comput Neurosci. 2018;12:33. doi:10.3389/fncom.2018.00033

20. Molina V, Bachiller A, De Luis R, et al. Topography of activation deficits in schizophrenia during P300 task related to cognition and structural connectivity. Eur Arch Psychiatry Clin Neurosci. 2019;269(4):419–428. doi:10.1007/s00406-018-0877-3

21. Bachiller A, Romero S, Molina V, et al. Auditory P3a and P3b neural generators in schizophrenia: An adaptive sLORETA P300 localization approach. Schizophr Res. 2015;169(1-3):318–325. doi:10.1016/j.schres.2015.09.028

22. Krčmář L, Jäger I, Boudriot E, et al. The multimodal Munich Clinical Deep Phenotyping study to bridge the translational gap in severe mental illness treatment research. Front Psychiatry. 2023;14:1179811. doi:10.3389/fpsyt.2023.1179811

23. Kalman JL, Burkhardt G, Adorjan K, et al. Biobanking in everyday clinical practice in psychiatry— The Munich Mental Health Biobank. Front Psychiatry. 2022;13:934640. doi:10.3389/fpsyt.2022.934640

24. Sheehan DV, Lecrubier Y, Sheehan KH, et al. The Mini-International Neuropsychiatric Interview (M.I.N.I.): the development and validation of a structured diagnostic psychiatric interview for DSM-IV and ICD-10. J Clin Psychiatry. 1998;59(20):22–33.

25. Kay SR, Fiszbein A, Opler LA. The Positive and Negative Syndrome Scale (PANSS) for Schizophrenia. Schizophr Bull. 1987;13(2):261–276. doi:10.1093/schbul/13.2.261

26. Keefe RSE, Goldberg TE, Harvey PD, Gold JM, Poe MP, Coughenour. The Brief Assessment of Cognition in Schizophrenia: reliability, sensitivity, and comparison with a standard neurocognitive battery. Schizophr Res. 2004;68(2-3):283–297. doi:10.1016/j.schres.2003.09.011

27. Adams RA, Pinotsis D, Tsirlis K, et al. Computational Modeling of Electroencephalography and Functional Magnetic Resonance Imaging Paradigms Indicates a Consistent Loss of Pyramidal Cell Synaptic Gain in Schizophrenia. Biol Psychiatry. 2022;91(2):202–215. doi:10.1016/j.biopsych.2021.07.024

28. Delorme A, Makeig S. EEGLAB: an open source toolbox for analysis of single-trial EEG dynamics including independent component analysis. J Neurosci Methods. 2004;134(1):9–21. doi:10.1016/j.jneumeth.2003.10.009

29. Gramfort A. MEG and EEG data analysis with MNE-Python. Front Neurosci. 2013;7. doi:10.3389/fnins.2013.00267

30. Andreychenko A, Boer VO, Arteaga De Castro CS, Luijten PR, Klomp DWJ. Efficient spectral editing at 7 T: GABA detection with MEGA-sLASER. Magn Reson Med. 2012;68(4):1018–1025. doi:10.1002/mrm.24131

31. Oeltzschner G, Zöllner HJ, Hui SCN, et al. Osprey: Open-source processing, reconstruction & estimation of magnetic resonance spectroscopy data. J Neurosci Methods. 2020;343:108827. doi:10.1016/j.jneumeth.2020.108827

32. Provencher SW. Estimation of metabolite concentrations from localized *in vivo* proton NMR spectra. Magn Reson Med. 1993;30(6):672–679. doi:10.1002/mrm.1910300604

33. John NA, Solanky BS, De Angelis F, et al. Longitudinal Metabolite Changes in Progressive Multiple Sclerosis: A Study of 3 Potential Neuroprotective Treatments. J Magn Reson Imaging. 2024;59(6):2192–2201. doi:10.1002/jmri.29017

34. Hall P. The Bootstrap and Edgeworth Expansion. Springer New York; 1992. doi:10.1007/978-1-4612-4384-7

35. Thulin M. Modern Statistics with R: From Wrangling and Exploring Data to Inference and Predictive Modelling. Second edition. CRC Press, Taylor & Francis Group; 2024.

36. Funatogawa I, Funatogawa T. Analysis of covariance with pre-treatment measurements in randomized trials: Comparison of equal and unequal slopes. Biom J. 2011;53(5):810–821. doi:10.1002/bimj.201100065

37. Kegeles LS, Mao X, Stanford AD. Elevated Prefrontal Cortex γ-Aminobutyric Acid and Glutamate-Glutamine Levels in Schizophrenia Measured In Vivo With Proton Magnetic Resonance Spectroscopy. Arch Gen Psychiatry. 2012;69(5):449. doi:10.1001/archgenpsychiatry.2011.1519

38. De Jonge JC, Vinkers CH, Hulshoff Pol HE, Marsman A. GABAergic Mechanisms in Schizophrenia: Linking Postmortem and In Vivo Studies. Front Psychiatry. 2017;8:118. doi:10.3389/fpsyt.2017.00118

39. Reinsel RA, Veselis RA, Heino R, Miodownik S, Alagesan R, Bedford RF. Effect of Midazolam on the Auditory Event-Related Potential: Measures of Selective Attention. Anesth Analg. 1991;73(5):612–618. doi:10.1213/00000539-199111000-00017

40. Rockstroh B, Elbert T, Lutzenberger W, Altenmüller E. Effects of the anticonvulsant benzodiazepine clonazepam on event-related brain potentials in humans. Electroencephalogr Clin Neurophysiol. 1991;78(2):142–149. doi:10.1016/0013-4694(91)90114-J

41. Watson TD, Petrakis IL, Edgecombe J, Perrino A, Krystal JH, Mathalon DH. Modulation of the cortical processing of novel and target stimuli by drugs affecting glutamate and GABA neurotransmission. Int J Neuropsychopharmacol. 2009;12(03):357. doi:10.1017/S1461145708009334

42. Smucny J, Dienel SJ, Lewis DA, Carter CS. Mechanisms underlying dorsolateral prefrontal cortex contributions to cognitive dysfunction in schizophrenia. Neuropsychopharmacology. 2022;47(1):292–308. doi:10.1038/s41386-021-01089-0

43. Murray JD, Anticevic A, Gancsos M, et al. Linking Microcircuit Dysfunction to Cognitive Impairment: Effects of Disinhibition Associated with Schizophrenia in a Cortical Working Memory Model. Cereb Cortex. 2014;24(4):859–872. doi:10.1093/cercor/bhs370

44. Hoftman GD, Datta D, Lewis DA. Layer 3 Excitatory and Inhibitory Circuitry in the Prefrontal Cortex: Developmental Trajectories and Alterations in Schizophrenia. Biol Psychiatry. 2017;81(10):862–873. doi:10.1016/j.biopsych.2016.05.022

45. Cen H, Xu J, Yang Z, et al. Neurochemical and brain functional changes in the ventromedial prefrontal cortex of first-episode psychosis patients: A combined functional magnetic resonance imaging— proton magnetic resonance spectroscopy study. Aust N Z J Psychiatry. 2020;54(5):519–527. doi:10.1177/0004867419898520

46. De La Fuente-Sandoval C, Reyes-Madrigal F, Mao X, et al. Cortico-Striatal GABAergic and Glutamatergic Dysregulations in Subjects at Ultra-High Risk for Psychosis Investigated with Proton Magnetic Resonance Spectroscopy. Int J Neuropsychopharmacol. 2016;19(3):pyv105. doi:10.1093/ijnp/pyv105

47. Reyes-Madrigal F, González-Manríquez L, Martínez De Velasco F, et al. Prefrontal γ-Aminobutyric Acid Levels in Never-Medicated Individuals With Chronic Schizophrenia. JAMA Psychiatry. 2023;80(10):1075. doi:10.1001/jamapsychiatry.2023.3157

48. Yang Z, Zhu Y, Song Z, et al. Comparison of the density of gamma-aminobutyric acid in the ventromedial prefrontal cortex of patients with first-episode psychosis and healthy controls. Shanghai Arch Psychiatry. 2015;27(6):341–347. doi:10.11919/j.issn.1002-0829.215130

49. Öngür D, Prescot AP, McCarthy J, Cohen BM, Renshaw PF. Elevated Gamma-Aminobutyric Acid Levels in Chronic Schizophrenia. Biol Psychiatry. 2010;68(7):667–670. doi:10.1016/j.biopsych.2010.05.016

50. De La Fuente-Sandoval C, Reyes-Madrigal F, Mao X, et al. Prefrontal and Striatal Gamma-Aminobutyric Acid Levels and the Effect of Antipsychotic Treatment in First-Episode Psychosis Patients. Biol Psychiatry. 2018;83(6):475–483. doi:10.1016/j.biopsych.2017.09.028

51. Poels EMP, Kegeles LS, Kantrowitz JT, et al. Glutamatergic abnormalities in schizophrenia: A review of proton MRS findings. Schizophr Res. 2014;152(2-3):325–332. doi:10.1016/j.schres.2013.12.013

52. Cooper JA, Nuutinen MR, Lawlor VM, et al. Reduced adaptation of glutamatergic stress response is associated with pessimistic expectations in depression. Nat Commun. 2021;12(1):3166. doi:10.1038/s41467-021-23284-9

53. Schmaal L, Goudriaan AE, Van Der Meer J, Van Den Brink W, Veltman DJ. The association between cingulate cortex glutamate concentration and delay discounting is mediated by resting state functional connectivity. Brain Behav. 2012;2(5):553–562. doi:10.1002/brb3.74

54. Bojesen KB, Broberg BV, Fagerlund B, et al. Associations Between Cognitive Function and Levels of Glutamatergic Metabolites and Gamma-Aminobutyric Acid in Antipsychotic-Naïve Patients With Schizophrenia or Psychosis. Biol Psychiatry. 2021;89(3):278–287. doi:10.1016/j.biopsych.2020.06.027

## Supplemental References

1. Kay SR, Fiszbein A, Opler LA. The Positive and Negative Syndrome Scale (PANSS) for Schizophrenia. Schizophr Bull. 1987;13(2):261–276. doi:10.1093/schbul/13.2.261

2. Andreasen NC, Carpenter WT, Kane JM, Lasser RA, Marder SR, Weinberger DR. Remission in Schizophrenia: Proposed Criteria and Rationale for Consensus. Am J Psychiatry. 2005;162(3):441–449. doi:10.1176/appi.ajp.162.3.441

3. Leucht S, Samara M, Heres S, Davis JM. Dose Equivalents for Antipsychotic Drugs: The DDD Method: Table 1. Schizophr Bull. 2016;42(suppl 1):S90–S94. doi:10.1093/schbul/sbv167

4. Heatherton TF, Kozlowski LT, Frecker RC, Fagerstrom K. The Fagerström Test for Nicotine Dependence: a revision of the Fagerstrom Tolerance Questionnaire. Br J Addict. 1991;86(9):1119–1127. doi:10.1111/j.1360-0443.1991.tb01879.x

5. Keefe RSE, Goldberg TE, Harvey PD, Gold JM, Poe MP, Coughenour. The Brief Assessment of Cognition in Schizophrenia: reliability, sensitivity, and comparison with a standard neurocognitive battery. Schizophr Res. 2004;68(2-3):283–297. doi:10.1016/j.schres.2003.09.011

6. Sachs G, Winklbaur B, Jagsch R, Keefe RSE. Validation of the German Version of the Brief Assessment of Cognition in Schizophrenia (BACS) – Preliminary Results. Eur Psychiatry. 2011;26(2):74–77. doi:10.1016/j.eurpsy.2009.10.006

7. Winkler I, Haufe S, Tangermann M. Automatic Classification of Artifactual ICA-Components for Artifact Removal in EEG Signals. Behav Brain Funct. 2011;7(1):30. doi:10.1186/1744-9081-7-30

8. Fan L, Li H, Zhuo J, et al. The Human Brainnetome Atlas: A New Brain Atlas Based on Connectional Architecture. Cereb Cortex. 2016;26(8):3508–3526. doi:10.1093/cercor/bhw157

